# Long-distance migration was associated with increased prevalence of post-traumatic stress disorder in Syrian refugees

**DOI:** 10.1101/2021.07.09.21259930

**Authors:** Andreas Halgreen Eiset, Michaelangelo P. Aoun, Monica Stougaard, Annemarie Graa Gottlieb, Ramzi S. Haddad, Morten Frydenberg, Wadih J. Naja

## Abstract

**Background:** Refugees are forced migrants but there is a large variation in the distance that refugees cover and there is a paucity in the evidence of how this may affect refugees’ health and health care needs. We investigated the association between long-distance migration and post-traumatic stress disorder (PTSD), a serious psychiatric disorder associated with deteriorating mental and somatic health.

**Methods:** Included from 2016-2019 were 712 adult Syrian refugees and asylum seekers in Lebanon and Denmark arrived up to 12 months prior. PTSD was assessed using the Harvard Trauma Questionnaire and the estimate of association was obtained by multiply imputing missing data and adjusting for confounding by propensity score-weighting with covariates age, sex, socioeconomic status, trauma experience and WHO-5-score, reporting the bootstrap 95-percentile confidence interval (95% CI). Additionally, a number of sensitivity analyses were performed.

**Results:** After multiply imputing missing data and propensity score-weighted adjustment for confounding, migration to Denmark instead of Lebanon was associated with an increase in PTSD prevalence of 9 percentage point (95% CI [-1; 19] percentage point).

**Discussion:** We found that long-distance migration was associated with an increase in the prevalence of PTSD suggesting that long-distance migration may be a factor to consider when assessing refugees’ and asylum seekers’ health. Practitioners should consider “long-distance migration” in refugee health screenings and in particular when assessing the risk of post-traumatic stress disorder. Future research should be designed to ultimately lead to studies of relevant interventions to lower the risk of post-traumatic stress disorder in refugees.

## Introduction

Refugees suffer from high risk of post-traumatic stress disorder (PTSD) [1] and are exposed to many risk-factors for PTSD before, during and after migration [2–4]. Risk factors for PTSD in the country of origin and in the host country has been examined in detail; however, the contribution of the migration itself has not received much attention in refugee health research. PTSD is a disorder that may develop after a traumatic event and is associated with significant loss of functioning and increased morbidity and mortality [5,6]. Symptoms of PTSD include intrusion (e.g. “flash-backs” and nightmares), avoidance and arousal, characterized by persistent perception of threat which may result in sleep deprivation, irritability and inability to concentrate [7,8]. Risk factors for developing PTSD comprise multiple trauma, female sex, being spouseless, ethnicity, lower educational status, comorbid mental disorders and type of trauma (for example intentional trauma, sexual assault etc.) [7]. Additionally, gene-environment interactions may play a key role [9]. Dysfunction of the amygdala (emotional learning and memory modulation) and medial prefrontal cortex (executive function) are key elements in PTSD and the disease is associated with epigenetic changes, increased blood levels of norepinephrine and decreased blood levels of glucocorticoids [7]. The life-time prevalence of PTSD in the general population is estimated to be around 5% for males and 10% for females with considerable variation according to the study population and diagnostic tools utilized [7,10]. In comparison, a recent meta-analysis produced a summary prevalence estimate for PTSD of 31% across all refugees [1] with little difference between males (29%) and females (34%) though with considerable variation between the country of origin, host countries and diagnostic tools; however, not taking migration history into account. The impact of migration and the role and measurement of “distance” (geographical, cultural, linguistic etc.) has been discussed at length for a number of outcomes [11,12] and long-distance migration has been associated with poorer self-rated health in the general population [13], hypothesizing a link with increasing difficulty in acculturation [14,15]. In the present study, we aimed at taking an initial step in assessing the effect of migration on refugees’ and asylum seekers’ health by comparing the prevalence of PTSD in newly arrived Syrian refugees and asylum seekers in Lebanon and Denmark.

## Methods

In a cross-sectional design with one-stage cluster randomized sampling between 2016 and 2019, newly arrived adult Syrian refugees were included in Lebanon and newly arrived Syrian asylum seekers were included in Denmark and assessed for PTSD using the Harvard Trauma Questionnaire (HTQ)-scale with cut-off score at 2.5. The implemented inclusion criteria were: (a) adult (≥18 years of age), (b) Syrian-born, (c) had left Syria after the onset of the ongoing civil-war (after February 2011), (d) arrived in Lebanon or Denmark less than 12 months prior to inclusion and (e) resident in the Lebanese settlements or Danish asylum seeker centres at the time of inclusion. Exclusion criteria were physical or mental illness that prevented participation. Variables to be collected to address confounding were decided upon *a priori* after discussions among the authors guided by graphical representations of our assumptions. See the Supplement 1-9 for a detailed description of the data collection instruments and the directed acyclic graph illustrating our assumptions about the relation of the variables. Also the study protocol is published and freely available [16].

Lebanon is a country of about 6 million people living in an area of nearly 10.000 km^2^—comparable in size to the two major Danish island (Zealand and Funen). Since 2015 Lebanon has stopped accepting asylum applications from Syrians seeking refuge in Lebanon. Nevertheless, being a close neighbour, in both geographical, linguistic and cultural distance, the influx of refugees from the Syrian war has endured. The majority live in structures ranging from regular (though dilapidated) structures to improvised tents clustered in informal gatherings scattered across Lebanon [17]. We stratified Lebanon into five regions based on geography, infrastructure and insights about local concerns (political, religious etc.). In each region, we collaborated with the local health service operators to draw up the sampling frame of gatherings and randomly decided on the inclusion sites, where all individuals eligible for inclusion were invited to participate (i.e. stratified cluster sampling [18,19]). See the Supplement 2 for details on the sampling frame. We obtained permission to include participants in all regions of Lebanon, that is, we had access to include participants from areas that are usually outside the reach of researchers due to “security considerations”. Should a chosen site be deemed unsafe at the scheduled time of inclusion we relocated to the closest setting deemed appropriate within the same region.

Denmark is a country of about 6 million people living in an area of about 50.000 km^2^. In Denmark, all eligible residents at a random sample of asylum centres were invited. The Danish asylum seeker centres are run by the Danish Immigration Services but outsourced to different operators. Several types of asylum seeker centres exist with the most common being the accommodation centre, to which the asylum seeker is allocated by random shortly after arrival and applying for asylum in Denmark. Of five available operators, three responded to the invitation, all accepting to participate in the study, representing a total of 11 accommodation asylum centres. Data-collection teams of two to four healthcare professionals were instructed during several pre-collection sessions in the practicalities and ethical considerations involved in the task: this encompassed obtaining written and informed consent, appropriate handling of confidential information, collection of the questionnaire data, the mental health self-rating scales, clinical measurements and the biological samples. At consent, the participant was instructed to independently complete a background questionnaire and mental health scales. The data collectors assisted those who could not read or write or had further questions regarding the study. Afterwards, the participant was invited to a semi-structured clinical interview and a short clinical exam of basic parameters (blood pressure, height, weight). Participants could at any time opt out of the data collection; if choosing to do so the participant was asked if she would care to provide a reason. Also, individuals who were invited but refused to participate were asked for a short non-structured interview including basic demographics and reason for refusal. The procedure was piloted in its entirety in Denmark including 10 participants in an asylum seeker centre, resulting in minor adjustments to the workflow. All relevant permissions were obtained prior to inclusion of any participant in both Lebanon and Denmark.

In the following we give a brief summary of the applied statistical methodology. Further details are given in the Supplement 3-9 and a thorough discussion of the statistical methodological considerations necessary to combine multiple imputation and propensity score-weighted analysis is provided in Eiset and Frydenberg [20]. Missing data was multiply imputed and confounding was address by propensity score-weighting including age, sex, socioeconomic status, experienced trauma and WHO-5-score in the model. The “standardised mortality ratio”-weights [21] was computed to estimate the weighted prevalence difference of PTSD in the population that fled to Denmark instead of Lebanon and presented with a 95-percentile confidence interval found by bootstrapping. A number of sensitivity analyses were carried out to assess the robustness of the estimated association between long-distance migration and PTSD. All data management, analysis and plots were done in R [22]. Further details are given in the Supplement 5 as well as directly in the R code available together with the *a priori* described analysis plan and a number of the exploratory plots from https://github.com/eiset/ARCH.

## Results

A total of 599 individuals were included in Lebanon and 113 were included in Denmark. Table 1 gives a statistical summary of the inclusion process and non-participation. The study populations differed on key variables such as sex (female: 73% in Lebanon, 47% in Denmark) and experience of violence (24% in Lebanon, 39% in Denmark) and there were missing values for all variables except long-distance migration. Figure 1 illustrates the missing proportion of key variables stratified on exposure status and Table 2 gives a detailed summary of the study population characteristics for both the observed realization and the imputed data set including missing for each covariates in the observed data.

**Table 1:** Summary of the inclusion in Denmark and Lebanon.

|  | **Denmark**<br>**n(%)** | **Lebanon**<br>**n(%)** |
| --- | --- | --- |
| Source population | 613<sup>a</sup> | 1,011,366<sup>a</sup> |
| Examined for eligibility | 124 | 641 |
| Excluded | 3 | 11 |
| Invited | 121 (100%) | 630 (100%) |
| Refused to participate |  |  |
| Total | 8 (7%) | 31 (5%) |
| Sex, female | 2 | 5<sup>b</sup> |
| Age<sup>c</sup> | All between 20 and 30 years old | All younger than 50 years old |
| Reasons<sup>d</sup> |  |  |
| Mistrust | 5 | 1 |
| No time | 2 | 4 |
| No personal gain | 0 | 2 |
| No answer | 1 | 24 |
| Included (participation rate) | 113 (93%) | 599 (95%) |

<sup>a</sup>The number given for the source population represents a maximum and include for example children. It was not possible to obtain a more precise estimate of the study population. <sup>b</sup>Sex was only recorded for 8 non-participants in Lebanon. <sup>c</sup>Few non-participants provided age (two in Denmark and two in Lebanon), thus a best guess is provided. <sup>d</sup>Reasons provided from the non-participants, aggregated into smallest possible number of strata.

**Figure 1:**
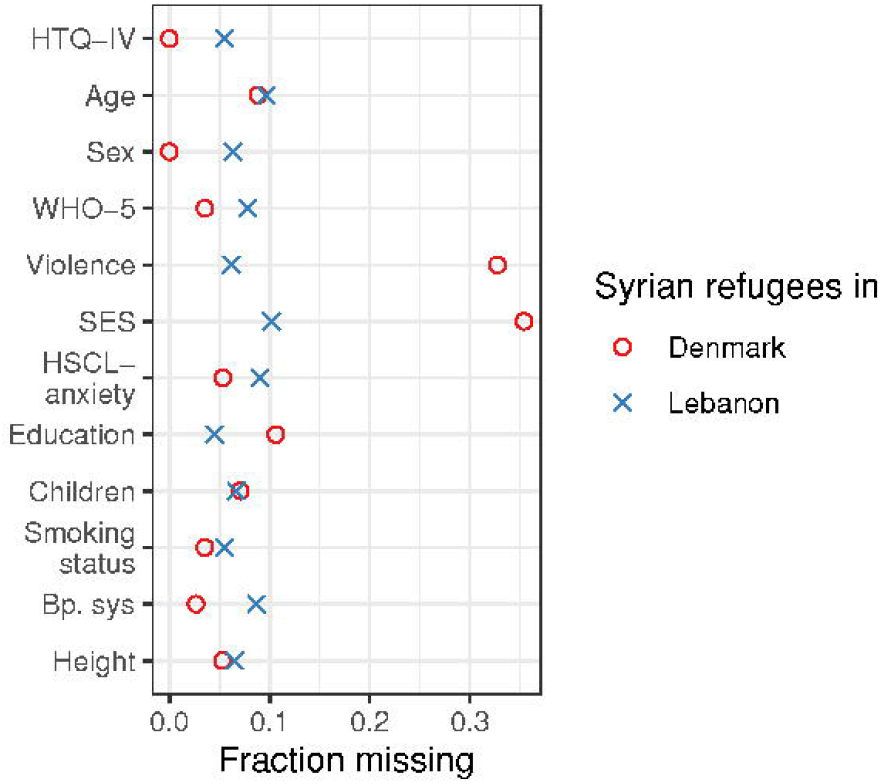
Missing fraction plot. Variables included in the propensity score and multiple imputation models for estimating the association between long-distance migration and PTSD among Syrian refugees in Lebanon and Denmark. Abbreviations: HTQ-IV, Harvard Trauma Questionnaire part IV; WHO-5, WHO-5 Mental-health scale; Violence, exposure (directly or indirectly) to violence; HSCL-anxeity, Hopkins Symptom Check List, anxiety part; Bp.sys, systolic blood pressure.

**Table 2:** Comparison of summary statistics of key variables in observed (with missing) and imputed data set stratified on exposure group.

|  | Observed data |  | Imputed data |  |
| --- | --- | --- | --- | --- |
|  | Lebanon<br>(n=599) | Denmark<br>(n=113) | Lebanon<br>(n=599) | Denmark<br>(n=113) |
| Age - years old | 35 (20) | 30 (12.5) | 35 (20) | 30 (12.9) |
| Missing | n=58 (9.7%) | n=10 (8.9%) | NA | NA |
| Sex |  |  |  |  |
| Female | 72.7% | 46.9% | 72.7% | 46.9% |
| Missing | n=38 (6.3%) | n=0 | NA | NA |
| WHO-5-score | 24 (32) | 24 (48) | 24 (32) | 24 (48) |
| Missing | n=47 (7.9%) | n=4 (3.5%) | NA | NA |
| Experienced violence |  |  |  |  |
| Yes | 23.8% | 39.5% | 23.8% | 39.7% |
| Missing | n=37 (6.2%) | n=37 (32.7%) | NA | NA |
| Socioeconomic status |  |  |  |  |
| Below average | 25.5% | 24.7% | 25.3% | 23.2% |
| On average | 63.4% | 50.7% | 63.4% | 50.0% |
| Above average<sup>a</sup> | 4.5% | 2.7% | 11.3%<sup>a</sup> | 26.8%<sup>a</sup> |
| Don't know/refuse to answer<sup>a</sup> | 6.7% | 21.9% |  |  |
| Missing | n=61 (10.2%) | n=40 (35.4%) |  |  |
| HTQ score | 2.56 (0.94) | 2.62 (1) | 2.56 (0.95) | 2.62 (1) |
| Missing | n=33 (5.5%) | n=0 | NA | NA |
| PTSD (HTQ score $\geq 2.5$) | | | | |
| Overall | 55.1% | 60.2% | 54.9% | 60.2% |
| Female | 59.4% | 66.0% | 59.1% | 66.0% |
| Male | 44.7% | 55.0% | 43.7% | 55.0% |
| Missing | n=33 (5.5%) | n=0 | NA | NA |

Categorical variables are represented as % of observations. Continuous variables are represented as median (interquartile range). Missing is given as n (%) of all included participants for the observed data
set. <sup>a</sup>In the substantive model, and thus in the multiple imputation, the strata “Above average” and “Don’t know/refuse to answer” was collapsed due to numerical problems in the multiple imputation.

The unadjusted prevalence of PTSD was higher in Denmark (60.2%) compared with Lebanon (55.1%). The prevalence difference of PTSD increased from 5.1 percentage point (95-percentile CI [-4.6; 15.0]) to 8.8 percentage point (95-percentile CI [-1.4; 18.6 percentage point]) after multiply imputing missing data and adjusting for confounding by propensity score-weighting. All sensitivity analyses produced estimates in the same direction and of the same magnitude, except when forcing all missing in the “Violence” variable to “Yes” in the study population included in Denmark, thus, grossly violating the missing-at-random assumption of multiple imputation. Table 3 presents the point estimate for the propensity score-weighted analysis and each of the sensitivity analysis accompanied by Figure 2 which includes different types of bootstrap confidence intervals.

**Table 3:** Estimated prevalence difference in crude, propensity score analysis and sensitivity analysis.

|  | Prevalence difference;<br>point estimate [95% CI]<sup>a</sup> |
| --- | --- |
| Multiply imputed and PS-weighted analysis | 8.8 [-1.4; 18.6]<sup>b</sup> |
| Complete case analyses |  |
| Crude | 5.1 [-4.6; 15.0]<sup>b</sup> |
| PS-weighted | 5.1 [-5.0; 16.1]<sup>b</sup> |
| Sensitivity analysis<sup>c</sup> |  |
| Alternative PS model | 8.0 [-1.4; 17.8] |
| HTQ score threshold for PTSD: 2.3 | 10.6 [0.6; 18.3] |
| HTQ score threshold for PTSD: 2.7 | 7.2 [-3.1; 16.7] |
| Missing in “SES” in Lebanon are forced to “Don’t know/refuse to answer” | 9.5 [-0.1; 19.6] |
| Missing in “Violence” in Denmark are forced to “Yes” | 4.2 [-5.4; 14.8] |

Abbreviations: CI, confidence interval; HTQ, Harvard Trauma Questionnaire; PS, propensity score; SES, socio-economic status. <sup>a</sup>All numbers are percentage point. The 95-percentile confidence interval is used for all estimates; <sup>b</sup>Based on 999 bootstrap replications, otherwise based on 250 bootstrap replications. <sup>c</sup>Multiply imputed and PS-weighted. Also see Figure 2.

**Figure 2:**
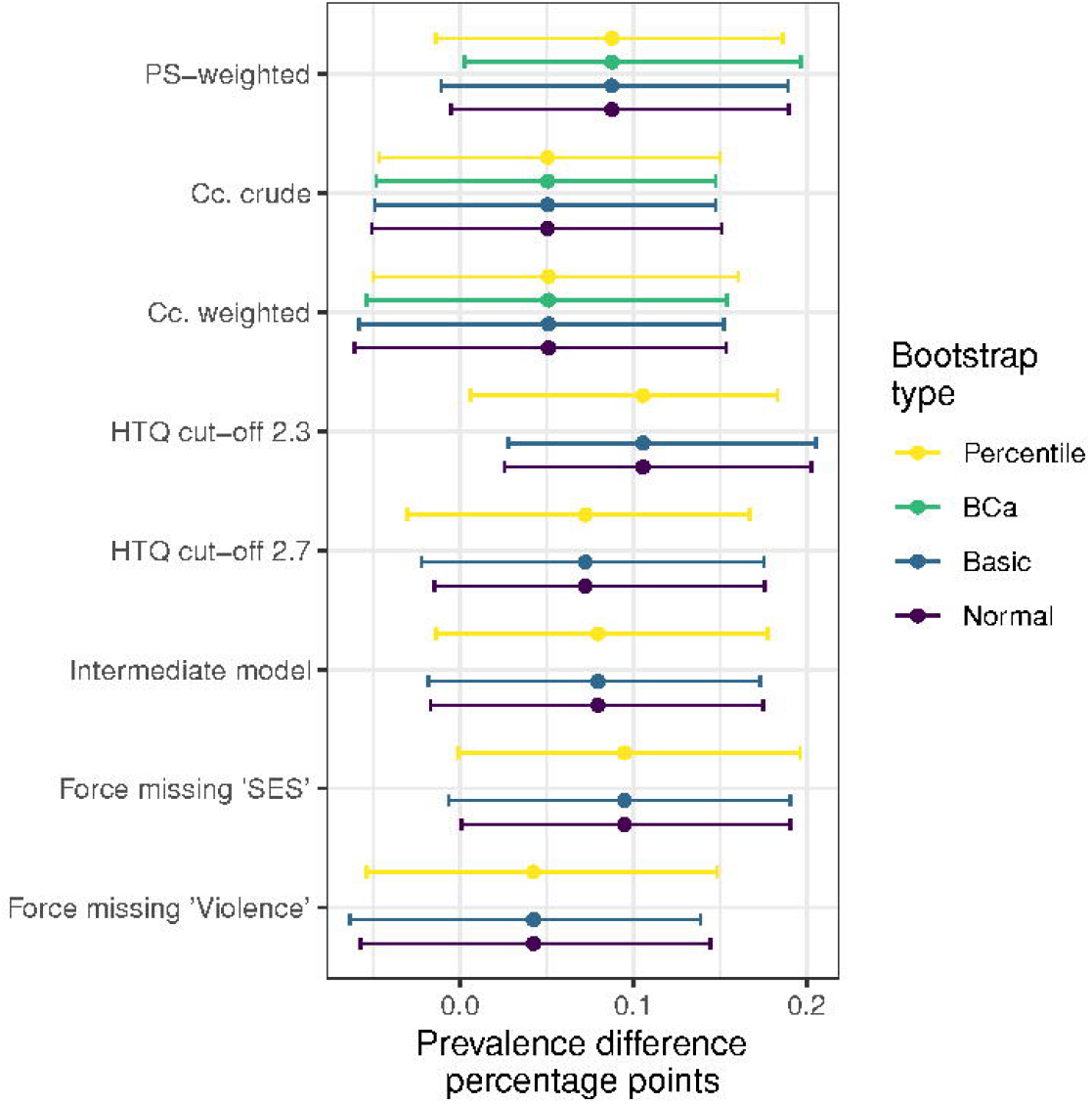
Estimate with different bootstrap confidence interval types. Abbreviations: BCa, bias-corrected and accelerated bootstrap CI; CI, confidence interval; PS, propensity score; Cc, complete case; HTQ, Harvard Trauma Questionnaire part IV; SES, socioeconomic status; Violence, exposure (directly or indirectly) to violence. From the top: the propensity score-weighted (“substantive model”) and the two complete case (no imputation) estimates with 999 bootstrap replications. The five sensitivity analysis are at the bottom, each with 250 bootstrap replications (and thus no BCa 95% CI).

## Discussion

This study aimed at estimating the association between long-distance migration and prevalence of PTSD. The estimate of association corresponded to 87 additional cases of PTSD for every 1000 Syrian refugees that migrated to Denmark instead of Lebanon. The 95% confidence interval indicate that, under the model, the estimate may be as high as 190 additional cases or as low as 4 cases less per 1000 refugees, thus, the estimate did not reach statistical significance, which may be ascribed to the relatively low number of participants. The targeted sample size was not reached neither in Lebanon nor in Denmark due to structural and political obstacles during the data collection.

To the best of the authors’ knowledge this is the first investigation of the association of long-distance migration and PTSD in a refugee population. A recent study [23] of Syrian refugees in Lebanon and Norway found a prevalence of PTSD (assessed as HTQ-score ≥ 2.5) of 4% in Lebanon and 7% in Norway. The study conflated heterogeneous groups for example asylum-seekers and refugees and participants with short and long stay in the host country prior to inclusion in the study. The study found that “Host country, Norway” was associated with an average increase in HTQ-score of 3% (95% CI [-5%; 12%]) when adjusting for age and sex; however, did not provide estimates of how this translated into absolute number of participants with PTSD. With a mean HTQ-score of 1.49 in the study population in Lebanon the number of additional cases in Norway due to “host country” would be marginal. The underlying cause for the observed increase in PTSD prevalence in Syrian refugees and asylum seekers after long-distance migration has never been investigated. In our study, we approach this by minimized the effect of living in the host country, thus, “long-distance migration” represented the migration process itself (travelling through Europe). We found a high prevalence of PTSD and an increased prevalence difference compared with the aforementioned study. Some studies hypothesise that great distance between populations is associated with difficulty acculturating which in turn is associated with worse health outcomes, including mental health [13,15,24]. The presented study is a first step in examining this in a refugee population.

The prevalence estimates of PTSD was high in both Lebanon and Denmark compared with previous studies of Syrian refugees and across different study designs and utilized instruments for assessing PTSD [1,25–27]. In the present study, we utilized the widely used HTQ-score [28] which is considered valid across multiple settings and diagnostic classification systems [29] although the specific cut-off has been debated [30]. The sensitivity analysis showed little change in the estimate of association including when changing the HTQ-score cut-off. An effort was made to make the data collection in Lebanon and Denmark as comparable as possible; however, differences may exist introducing information bias. For example, differential misclassification cannot be ruled out, if, during data collection in Denmark, doubt remained among the participants whether specific answers would affect their asylum application. This may explain (some of) the difference in prevalence of PTSD in Lebanon and Denmark; however, judged from extensive exploratory plotting of the covariates, there was no difference between the exposure groups in the association of the reported HTQ-score and WHO-5 (general mental well-being) or depression- or anxiety-scores, thus, the participants would have had to be very consistent in their misclassification of own mental health. The proportion that refused to answer, or did not know what to answer, in the question on socioeconomic status was high and differed between the study population in Denmark (22%) and in Lebanon (7%). After discussing this variable with Syrian natives, we speculate that “socioeconomic status” was not defined in sufficient detail for all participants to be able to reflect on the question; however, we do not have an explanation for the difference in proportion of missing between Lebanon and Denmark. We consider the risk of selection bias low given the randomized sampling design and very high participation rates. It was, however, not possible to retrieve or create a precise sampling frame in Lebanon and thus, it was based on best information available at the time, knowing that refugees frequently relocate and there is little registration of individuals outside the formalized refugee camps.

In the present study, the interpretation of the association between long-distance migration and PTSD is of course limited by the cross-sectional study design. Most importantly, the time-order may be reversed so that instead of long-distance migration leading to higher risk of PTSD it may be that PTSD affect the “risk” of undertaking long-distance migration. A positive association between long-distance migration and PTSD was found, thus, a reversal of the time order would mean that individuals with PTSD are more likely to undertake long-distance migration. Although this cannot be entirely rejected, taking into account the classical symptoms of PTSD such as difficulties in concentrating and planning, this interpretation of the result seems far less likely than our stated hypothesis: that undertaking an, at times, hazardous travel and living with uncertainty about the near future, increases the risk of PTSD.

We addressed confounding by propensity score-weighting and obtained balance on all covariates judged from our pre-specified criteria. To account for confounding we included covariates in the propensity score model according to what the authors believe is the current best evidence [7]; however, this does not alleviate us of speculations of entirely other covariates that may change the estimate. Furthermore, the collapsing of two levels of the socioeconomic status covariate diminishes the information this variable contains and thus may be a source for residual confounding in both the multiple imputation and in the propensity score weighted analysis. The missing mechanism was “everywhere missing-at-random” allowing for frequentist inference from the multiply imputed data. In the Supplement 4 we provide examples of the discussion necessary for each partly observed variable to deem the missingness mechanism “everywhere missing-at-random”.

Future research should be designed to determine if the association is transportable to other groups of refugees and to investigate the underlying factors contributing to the association, for example examine the different measures of “distance” (cultural, geographical etc.), the different types of trauma refugees may experience during migration (poor shelters, starvation, interpersonal violence etc.), means of transportation during the migration and the (health) reception in the host country. We note that the direction of the association was consistent across different thresholds for PTSD (effectively lowering and raising the estimated prevalence for PTSD in the study population), different propensity score models and when imposing violation of underlying assumptions of multiple imputation; the strength was also relatively consistent. We hypothesize that the association will be reproducible at least in comparable populations and settings—for example newly arrived adult refugees from a Middle Eastern country to a Western European country. Also, the association between long-distance migration and other health outcomes, for example other mental health issues, is of great interest. The presented study is an initial analysis of the association between long-distance migration and PTSD and provides the foundation upon which future studies could build.

## Data Availability

The individual-level data cannot be shared because it contains sensitive information and its distribution would violate the agreement under which the data was provided as well as Danish law. The computing code for all parts of the data cleaning, analysis, and plotting is publicly available from the first author's Github repository: https://github.com/eiset/ARCH. This repository also contains all the exploratory plots produced.

## Author contributions

Conceptualization: AHE; Methodology: AHE, MF; Formal analysis and investigation: AHE, MF; Writing - original draft preparation: AHE; Writing - review and editing: AHE, MPA, MF, AGG, MS, WJN, RSH; Funding acquisition: AHE; Resources: AHE, MPA, WJN, RSH, AGG, MS; Supervision: MF, RSH, MS, AGG, WJN.

## Acknowledgements

The authors would like to thank the following for their invaluable help in the data collection, in no particular order: Dr. Fayez Saadaldin, Margrethe Nielsen, Susanne Løgsted, Rebekka Pedersen, Sine Søe Romedahl, Hanne Jeppesen, Dr. Chahid Farah, Dr. Fadi Nader, Dr. Malak Rammal, Dr. Mayssaloun Khairallah, Dr. Sultana Baydoun, Dr. Tanios Dagher, Dr. Maibritt Provstgaard, Maja Nielsen, Dr. Arendse Lykke Loua, Dr. Cecilie Blenstrup Patsche, Dr. Bjørg Olsen, Dr. Camilla Kjersgaard, Hawa-Idil Harakow.

## Ethics and research committee approval

All relevant permissions and approvals were obtained prior to inclusion of any participant in Lebanon and Denmark including from the Lebanese Ministry of Public Health (2018/4/38918), the Ethics Committee of Mount Lebanon Hospital (PSY-2018-005), the Ethics Committee of the Lebanese University (CUMEB/D 144/232018) and the Danish Data Protection Agency (J.nr. 2015-41-4500). The Danish National Committee on Health Research Ethics waived the right to assess the project (F 257/2015).

