## Supplement for "Long-distance migration was associated with increased prevalence of post-traumatic stress disorder in Syrian refugees"

Michaelangelo P. Aoun[2], MD

Monica Stougaard[1], PhD

Annemarie Graa Gottlieb[1], PhD

Ramzi S. Haddad[2], MD

Morten Frydenberg[3], PhD

Wadih J. Naja[2,4], MD

[1]Department of Affective Disorders, Aarhus University Hospital–Psychiatry, Aarhus, Denmark

[2]Faculty of Medical Sciences, Lebanese University, Beirut, Lebanon

[3]Consultant biostatistician, Trige, Denmark

[4]King Hussein Cancer Center, Amman, Jordan

*Corresponding author

Andreas Halgreen Eiset

The aim of the study was to estimate the association between long-distance migration and prevalence of post-traumatic stress disorder (PTSD) in newly arrived adult Syrian asylum seekers in Denmark and newly arrived adult Syrian refugees in Lebanon.

We defined the exposure, “long-distance migration”, as having migrated to Denmark instead of Lebanon. The outcome, PTSD, was assessed using the The Harvard Trauma Questionnaire (HTQ) which was developed in 1992 among others to assess symptoms of PTSD according to the DSM-IV criteria (part IV) [1]. The The Harvard Trauma Questionnaire part-IV (HTQ) has been validated in multiple settings, languages and populations [2–4]. Each of the initial 16 items, corresponding to the 17 DSM-IV post-traumatic stress disorder symptoms, is scored on an ordinal scale from “not at all” (item score = 1) to “extremely” (item score = 4). To obtain the DSM-IV score, the mean is calculated and a cut-off score of 2.5 or more is often taken to indicate PTSD. It is debated whether the cut-off score transportable in cross-cultural settings [5]. The validated Arabic 2007 version [2] was utilised. The HTQ scale is available for purchase from http://hprt-cambridge.org/screening/harvard-trauma-questionnaire/

We collected information on “general mental well-being” using the WHO-5 scale which is freely available from https://www.psykiatri-regionh.dk/who-5/who-5-questionnaires/Pages/default.aspx Further details are given in the study protocol [6].

Below is a subset of the questionnaire developed to collect background information on participants after written and oral informed consent to participate. The questionnaire was developed in Danish, translated by a bilingual and bicultural translators: into Arabic (mother tongue Syrian Arabic), back translated by two translators (mother tongue was Arabic (other Arabic dialects than Syrian), each item was evaluated and minor corrections was implemented. The final translated questionnaire was approved by three Lebanese medical doctors (mother tongue Lebanese Arabic). The participants were allowed to opt out for parts of the data collection and continue with the rest.

Below, we reproduce parts of the questionnaire in English translated by the authors of the manuscript. This translation has never been used for data collection and has not been validated by a translator.

The analysis plan was specified a priori and is supplied at https://github.com/eiset/ARCH and the applied methodology is discussed in detail in Eiset and Frydenberg [7].

### Supplement 1: The background questionnaire

1. About you and your family

1.a Your sex (circle correct answer): male female

1.b Your age (in years):

1.c Your marital status (circle correct answer): unmarried married widowed other

1.d How many children do you have (circle correct answer): 0 1 2 3 4 5or more

2. About your education

2.a What is your highest obtained educational level (circle correct answer): none elementary gymnasium higher

2.b How many years of education have you had (in years):

2c How would you rank your social status before leaving Syria (circle correct answer): below average on average above average do not know

3. About your health

3.a Do you smoke tobacco (circle correct answer): yes no, but previously no, never

4. About your migration

4.a On what date did you leave Syria (year, month, day, as precisely as you remember):

4.b On what date did you arrive in [Lebanon/Denmark] (year, month, day, as precisely as you remember):

4.c What means of travel did you use to get from Syria to [Lebanon/Denmark] (circle all applicable): airplane car bus train boat walked long distances other

4.d During your travel, did you experience violence or abuse (circle all applicable): yes, first hand yes, second hand no

### Supplement 2: Sample size and strata

We aimed at a sample size of 1100 participants in Lebanon and 220 participants in Denmark. In an independent two-group design without continuity correction this allows us to show a statistically significant difference between a prevalence of 5% and 12% (representing the cut-off under which the outcome was deemed “rare” and the cut-off above which the outcome was deemed “not uncommon”) with a statistical significance level of 5%, power of 80% and survey design effect of one assuming random allocation to clusters [8]. In Lebanon, proportional allocation in each region was secured by estimates of the source population using UNHCR statistics [9,10] resulting in sample size aims in each strata as presented in the “Table of sample size aim” below.

**Table of sample size aim for strata with proportional allocation**

| | **Estimated source population N=1,011,366 (100%)** | **Sample size aim N=1100 (100%)** |
|---|---|---|
| Bekaa valley | 362,069 (35.8%) | 395 (35.9%) |
| North | 261,978 (25.9%) | 286 (26.0%) |
| Mount Lebanon | 164,898 (16.3%) | 186 (16.9%) |
| South | 121,057 (12.0%) | 133 (12.1%) |
| Beirut | 101,364 (10.0%) | 100 (9.1%) |

*For each stratum, the estimated source population and the aimed sample size, both with percentage of the reference population. The estimated source population constitute the entire Syrian refugee population, including for example children, in the given strata at the time of initiating data collection in Lebanon. It was not possible to obtain an estimate of the source population that fulfilled the inclusion criteria.*

### Supplement 3: The propensity score-weighted estimate of association

Variables to include to control confounding and potential confounding were age, sex, mental health co-morbidity, experience of violence and SES. Three propensity score models with increasing complexity was proposed (see “Table of Covariates” below) and covariate balance in the exposure groups was assessed with no truncation, truncation at 1st and 99th percentile and at 5th and 95th percentile (Figure S2). The simplest model with the least amount of truncation that obtained acceptable balance, defined as an absolute mean difference less than or equal to 0.10 for all covariate, was used for the propensity score analysis.

**Table of Covariates in the three proposed propensity score models and how they enter. "Long-distance migration" is the response in all models**

| | Enter as | Model 1: "simple" | Model 2: "intermediate" | Model 3: "complex" |
|---|---|---|---|---|
| Age | 3-knots restricted cubic spline | x | x | x |
| Sex | Dichotomous | x | x | x |
| WHO-5 | 3-knots restricted cubic spline | x | x | x |
| Violence | Dichotomous | x | x | x |
| SES | 3-level ordinal | x | x | x |
| Sex & age | Interaction, no transformation | | x | x |
| Sex & WHO-5 | Interaction, no transformation | | | x |
| Age & WHO-5 | Interaction, no transformation | | | x |

*Abbreviations: Interact., interaction term; WHO-5, the WHO-5 quality of life scale; SES, socio-economic status; Violence, experience (direct or indirect) of violence during migration (from Eiset and Frydenberg [7]).*

From Figure S2 it is evident that the simple model with truncation at 1st and 99th percentile obtained balance on all covariates. For example, for the sex variable, in Lebanon approximately 73% of participants were female, whereas this was the case for 47% of the participants included in Denmark, resulting in a difference of 26 percentage point. In the re-weighted "pseudo-population" the mean difference was reduced to 4 percentage point.

### Supplement 4: The multiple imputation modeling of missing data

The SMC-FCS implementation of multiple imputation was utilized as detailed in Eiset and Frydenberg [7]. Here, the substantive model (here the propensity score model) entered separately and a "prediction model" (used here to describe a model to predict the value of the covariate in question given the other covariates and possible auxiliary variables) was specified for each partially observed covariate and any auxiliary variables. The extensive exploratory analysis including plots and tables are available from https://github.com/eiset/ARCH and the resulting "response-and-predictor matrix" and details of how each variable was treated is supplied in "The predictor matrix" below.

### The predictor matrix for the SMC-FCS multiple imputation

| | | Predictors | | | | | | | | | | | | | | | | | | | | | | | | |
|---|---|---|---|---|---|---|---|---|---|---|---|---|---|---|---|---|---|---|---|---|---|---|---|---|---|---|
| **Type** | **Response** | migr | age_log | age | age_sb | sex | sexFemale:age | who_sqrt | who | who_sb | who:sexFemale | who:age | viol | ses | ptsd | ptsd_sb | edu | child | smok | bp_log | bp | bp_sb | hgt_log | hgt | hgt_sb | mari |
| b | *migr* | | | | | | | | | | | | | | | | | | | | | | | | | |
| c | *age_log* | | | | | | | | | | | | | 1 | 1 | 1 | 1 | 1 | | | 1 | 1 | | | | 1 |
| pas | age | | 1 | | | | | | | | | | | | | | | | | | | | | | | |
| pas | age_sb | | | 1 | | | | | | | | | | | | | | | | | | | | | | |
| b | *sex* | | | | | | | | | | | | | | 1 | 1 | | | 1 | | 1 | 1 | | 1 | 1 | 1 |
| pas | sexFemale:age | | | 1 | | 1 | | | | | | | | | | | | | | | | | | | | |
| c | *who_sqrt* | | | 1 | 1 | 1 | 1 | | | | | | 1 | 1 | 1 | 1 | | | | | | | | | | |
| pas | who | | | | | | | 1 | | | | | | | | | | | | | | | | | | |
| pas | who_sb | | | | | | | | 1 | | | | | | | | | | | | | | | | | |
| pas | who:sexFemale | | | | | 1 | | | 1 | | | | | | | | | | | | | | | | | |
| pas | who:age | | | 1 | | | | | 1 | | | | | | | | | | | | | | | | | |
| b | *viol* | | | 1 | 1 | 1 | 1 | | 1 | 1 | 1 | | | | 1 | 1 | | | | | | | | | | |
| o | *ses* | | | 1 | 1 | 1 | 1 | | 1 | 1 | 1 | | | | 1 | 1 | | 1 | | | | | | | | 1 |
| c | *ptsd* | | | 1 | 1 | 1 | 1 | | 1 | 1 | 1 | 1 | 1 | 1 | | | | | | | | | | | | |
| pas | ptsd_sb | | | | | | | | | | | | | | 1 | | | | | | | | | | | |
| o | *edu* | | | 1 | 1 | 1 | 1 | | | | | | | 1 | | | | | | | | | | | | 1 |
| o | *child* | 1 | | 1 | 1 | 1 | 1 | | | | | | | 1 | | | 1 | | | | 1 | 1 | | | | 1 |
| b | *smok* | 1 | | | | 1 | | | | 1 | 1 | | 1 | | | | | | | | 1 | 1 | | 1 | 1 | |
| c | *bp_log* | | | 1 | 1 | 1 | 1 | | | | | | | | | | | | | | | | | | | |
| pas | bp | | | | | | | | | | | | | | | | | | | 1 | | | | | | |
| pas | bp_sb | | | | | | | | | | | | | | | | | | | | 1 | | | | | |
| c | *hgt_log* | | | | | 1 | | | | | | | | | 1 | 1 | | | 1 | | 1 | 1 | | | | |
| pas | hgt | | | | | | | | | | | | | | | | | | | | | | 1 | | | |
| pas | hgt_sb | | | | | | | | | | | | | | | | | | | | | | | 1 | | |
| u | *mari* | | | | | 1 | | | | | | | | | | | | 1 | | | | | | | | |

*The predictor matrix details how each partially observed variable was imputed. Horizontally the variables enter as the response, vertically the variables enter as the predictor when the corresponding cell is "1". The "type" column indicate how the variables should be modelled when entering as the response (b, binary; c, continuous; pas, passive; o, ordered/ordinal; u, unordered/nominal). Variables ending on "_sb" are modelled as restricted cubic spline (spline basis); ":" indicates interaction between the variables on each side. Variables in red are part of the substantive model (the propensity score model); variables in italic are actively imputed. From* [7].

An example of the discussion of missing data follows using the variables "Children" (i.e. the participant's number of offspring) and "Age" (i.e. the participant's age in years at time of inclusion). While Children is not in the substantive model (i.e. does not enter the propensity score model) and might be considered less important in this analysis, it holds valuable information on a very important variable: Age (Supplement 8). Therefore, it enters as an auxiliary variable in the prediction model for Age. Before doing so, the missing data in Children (Figure 1) must be imputed by specifying a prediction model with Children as the response ("The predictor matrix", above) and the validity of multiple imputation must be discussed for Children. In our analysis, the assumption of ignorable missingness mechanism requires that the missingness mechanism is at least "everywhere missing-at-random" [11], i.e. that any possible missingness pattern of Children is independent of the missing values in Children given the covariates and the observed values for Children (see elaborations in [12]). If it is more likely that an individual with no children did not answer the question and if no other variables are considered, then the missingness mechanism is at best "realized missing-at-random" (if all instances of Children = 0 are realized, which we cannot know). In the case of Children, among others low age seems to increase the probability of data being missing (Supplement 8). See "The predictor matrix" above for the other variables that were included as predictors in modeling Children based on subject matter knowledge and the exploratory analysis https://github.com/eiset/ARCH. Assuming independent and identical missingness mechanism, we considered the possible missingness patterns for one individual, and judged that the "everywhere missing-at-random" assumption was well approximated. Next, the assumption of correctly specified multiple imputation depends on whether the substantive model of interest is correctly specified and whether it is possible to build a multiple imputation model that captures the entirety of the data generating process. Again, this relies on subject matter insight, a thorough discussion of the proposed model and a deep understanding of the applied methods.

For the imputation of the partly observed variable Age, which is in the substantive model, we again rely on sufficient information being observed so that we may predict the missingness pattern and assume that the missingness mechanism is approximately "everywhere missing-at-random". A strong predictors of age is systolic blood pressure (Supplement 9). Systolic blood pressure not in our propensity score model, thus, it is an auxiliary variable. It has missing values (Figure 1) and so also needs imputing…

### Supplement 5: Sensitivity analysis

To evaluate the robustness of the results of the analysis, the following analyses were planned:

- Evaluating the model choice for the propensity score analysis using other propensity score models (Table S2) that obtained acceptable balance on all covariates.
- The cut-off value of the HTQ score to indicate PTSD was set to the "standard" ≥2.5. The estimates and corresponding 95% CI from models with cut-off values at ≥2.3 and ≥2.7 is supplied.
- Assessing the impact of non-ignorable missingness mechanism by forcing all missing in the "socio-economic status" covariate in participants from Lebanon to be "Do not know/refuse to answer" and forcing all missing in the "Experienced violence during migration" covariate in participants from Denmark to "Yes". In both scenarios all other missing were kept as imputed.

### R code and packages

All data management, analysis and plots were done in R [13] with heavy reliance on the "Tidyverse" packages [14] for data management and plots, "smcfcs" [15] for multiple imputation, "WeightIt" [16] for obtaining the propensity score weights and "boot" [17] for parallelized bootstrapping. The full list of utilized packages as well as the R code for analysis and plots is available from https://github.com/eiset/ARCH.

### Supplement 6: the DAG

**Directed acyclic graph: assumptions of the association between long-distance-migration and post-traumatic stress disorder.**

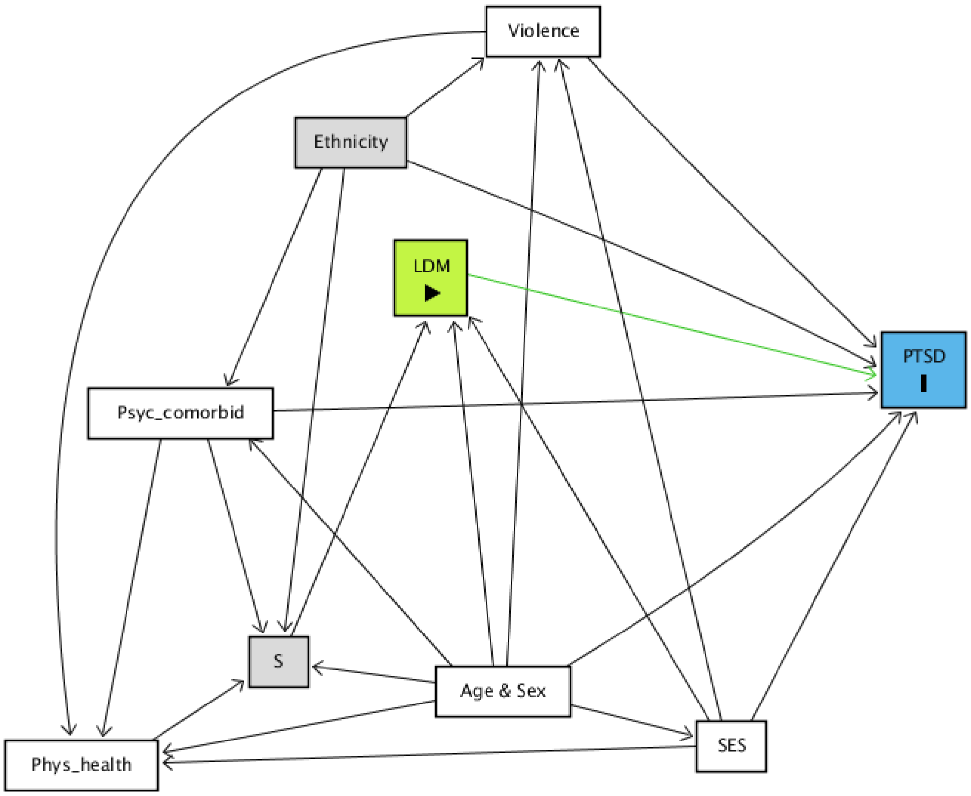

*Abbreviations: S, selection variable, variables going into this define the study population; LDM, long-distance migration (the exposure of interest); PTSD, post-traumatic stress disorder (the outcome of interest); SES, socio-economic status; Psyc_comorbid, psychiatric co-morbidity; Phys_health, physical health; Violence, exposure (directly or indirectly) to violence. Ethnicity is used to capture genetics background, physical phenotype, parental physical phenotype and cultural context. Likewise, age and sex is collapsed in this depiction after securing no loss of information. Colour code: Green, exposure; Blue, outcome; Grey, covariates that are controlled for by design; White, other covariates.*

### Supplement 7: Balance plot

**Balance plots for the three complexities of the propensity score model and the three cut-offs for weight truncation.**

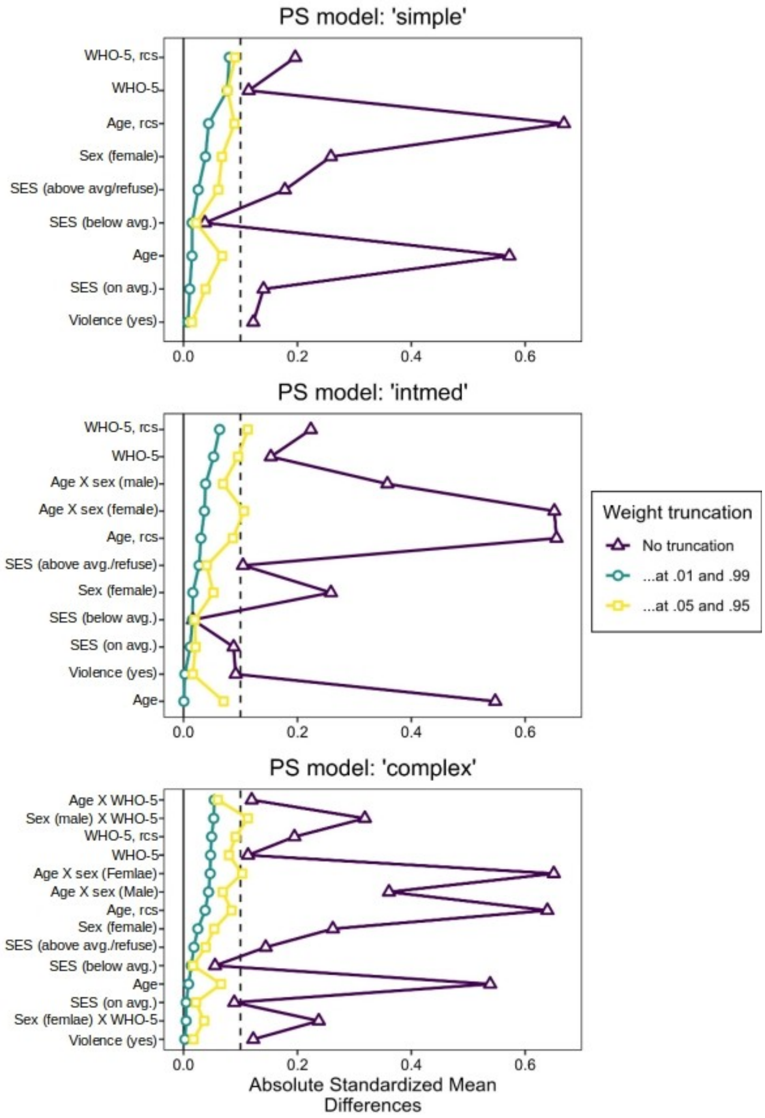

*Abbreviations: WHO5, WHO (five) Well-being Index; rcs, three-knots restricted cubic spline term; SES, socioeconomic status; avg, average; Violence, exposure (directly or indirectly) to violence; X, interaction term.*

#### Supplement 8: Plot of number of children against age

**Violin plot of number of children against the participant's age.**

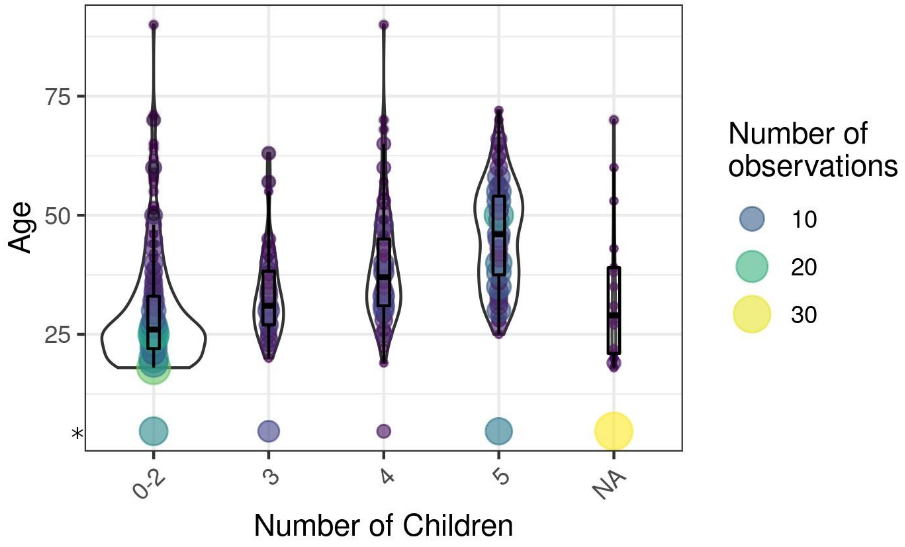

**Missing values in Age are plotted as (minimum value minus mean absolute deviation). The relative point size and colour gradient indicate the density of observations at each space. It is clear that the number of children is a fairly good predictor of age (e.g. in the case of 2 versus 3 children the "best guess" of the age of a participant with missing information on age would change from approximately 25 years old to approximately 33 years old).*

### Supplement 9: Plot of blood pressure against age

**Systolic blood pressure against the participant's age.**

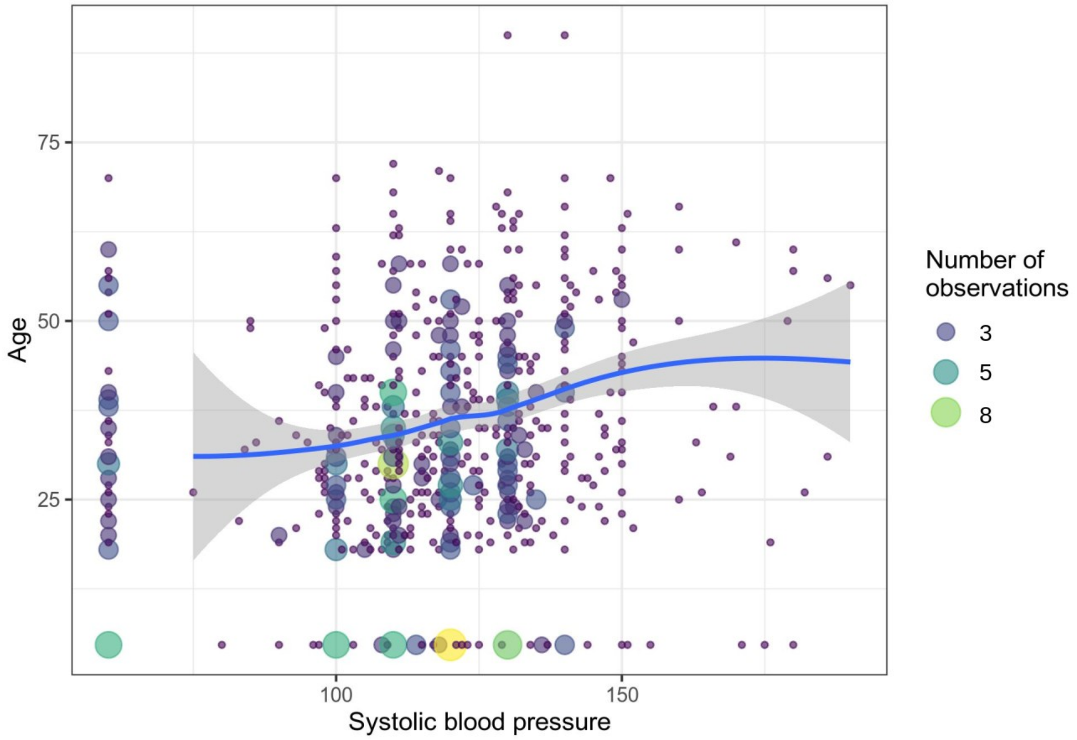

**Missing values are plotted as (minimum value minus mean absolute deviation). The relative point size and colour gradient indicate the density of observations at each space. It is clear that the blood pressure is a strong predictor of age (in accordance with subject matter knowledge).*
